# Prevalence of Manifestations of Afferent Baroreflex Failure Among Long-Term Survivors of Oropharyngeal Cancer

**DOI:** 10.1101/2025.09.08.25335364

**Authors:** Patrik J. Van-Bergen, Dhawal S. Bandrey, Juhee Song, Stephen P. Patin, Neeharika Prathapa, Gabrielle N. Mbagwu, Andres Hughes, Jennifer Shen, Mohamed A. Naser, Katherine A. Hutcheson, David I. Rosenthal, Amy C. Moreno, Elie Mouhayar, Anita Deswal, Clifton D. Fuller, Efstratios Koutroumpakis, OPC-SURVIVOR Program MD Anderson Head and Neck Radiation Oncology and Cardiovascular Working Group MD Anderson Head and Neck Cancer Symptom Working Group

## Abstract

**Background:** Afferent baroreflex failure (ABF) is an underrecognized but debilitating complication among head and neck cancer survivors, especially in oropharyngeal cancer (OPC), a malignancy with excellent prognosis. ABF is mainly caused by radiation therapy (RT), with neck surgery and some chemotherapies also contributing. It manifests as blood pressure lability, including severe hypertension or hypotension, syncope, and arrhythmias.

**Objectives:** To determine the prevalence and predictors of ABF-associated manifestations among OPC survivors treated with modern RT.

**Methods:** We retrospectively studied OPC patients treated with RT at a tertiary cancer center between 2016–2019. Clinical data were collected from RT initiation to last follow-up or death. ABF-associated manifestations included new or worsening hypertension, hypotension requiring intervention, arrhythmias, and syncope. Secondary endpoints included new or worsened carotid artery atherosclerosis, stenosis, transient ischemic attack (TIA), stroke, and all-cause mortality.

**Results:** Among 393 patients (88% men, 91% White, mean age 61±10 years), 9.4% developed hypertension, 5.3% hypotension, 3.8% syncope, and 3.3% arrhythmias over median 6.3-year follow-up. Overall, 19.1% developed at least one ABF-associated manifestations. New or worsened carotid atherosclerosis occurred in 38.9%, with 7.1% developing >50% stenosis and 2.3% experiencing TIA or stroke. Mortality was 21.4%. On cause-specific multivariable Cox analysis, older age (adjusted hazard ratio [aHR] 1.03; 95% confidence interval [CI] 1.01–1.06), valvular disease (aHR 2.85; CI 1.03–7.92), T4 cancer (aHR 1.90; CI 1.10–3.27), and platinum-taxane chemotherapy (aHR 1.86; CI 1.13–3.05) independently increased risk of ABF-associated manifestations.

**Conclusions:** Nearly 1 in 5 OPC survivors treated with RT develop ABF-associated manifestations, highlighting the need for early recognition and surveillance.

## Introduction

Radiation therapy (RT) remains a cornerstone in the treatment of oropharyngeal cancer (OPC), with recent advancements significantly enhancing survival outcomes over the past decade, as reflected by a rise in reported 5-year survival rates to approximately 85%.^1^ As survival rates have improved, focus has increasingly shifted toward understanding and mitigating the long-term complications of treatment. Among these, radiation-induced vascular injury—particularly carotid artery stenosis and its sequelae, such as transient ischemic attack (TIA) or stroke—represents a major source of morbidity.^2^ Another form of radiation-induced vascular toxicity, afferent baroreflex failure (ABF), remains less recognized, yet is potentially debilitating and clinically significant.^3,4^

ABF manifests as severe blood pressure lability, including episodes of severe hypertension or hypotension, recurrent syncope, and arrhythmias, leading to a substantial decline in quality of life.^5^ It typically manifests years after neck irradiation, often presenting insidiously but progressing to chronic and irreversible symptomatic sequelae once clinically apparent.^6^ Currently, there are no evidence-based therapies to prevent ABF and once the condition becomes clinically apparent, treatment options are limited. Pharmacologic approaches such as midodrine and fludrocortisone for hypotension, and centrally acting alpha-agonists for hypertension, have been used with limited efficacy.^7^ This therapeutic gap underscores the importance of early recognition and diagnosis. While no preventive strategies have been formally established, emerging studies suggest that agents like statins may hold potential in the prevention of radiation-induced vascular damage.^8^ These preliminary findings warrant further investigation and highlight the value of increased clinical awareness and proactive research into targeted interventions.

Although ABF can significantly impair quality of life, it remains underdiagnosed—largely due to its delayed and nonspecific clinical presentation, coupled with the absence of routine screening in oncology follow-up appointments.^7^ Autonomic function testing, required for definitive diagnosis, is rarely performed outside specialized centers, resulting in years-long diagnostic delays that limit opportunities for timely intervention and hinder the development of targeted therapies.^5,9^ Importantly, manifestations commonly seen in ABF—such as blood pressure lability, syncope, and arrhythmias—are not pathognomonic but are prevalent among cancer survivors.^10^ Recognizing these patterns can serve as a clinical signal to initiate ABF-specific testing in at-risk patients, particularly those with prior neck irradiation.^11^ Bridging this diagnostic gap is critical to improving clinical awareness, enabling timely referral and diagnosis, and laying the groundwork for the investigation of potential preventive strategies. ^12^

This study aims to characterize the relative prevalence of ABF-associated manifestations among long-term (>5-year) OPC survivors treated with modern RT and to identify patient- and treatment-related factors associated with their development. By systematically evaluating a well-defined cohort over extended follow-up, this analysis seeks to determine relative frequency and context in which these underrecognized symptoms arise, to inform improved detection and post-treatment care strategies in this high-risk population, as well as defining cohorts for future prospective research surveillance and intervention studies.

## Methods

### Patient Population and Data Collection

Cases of adult (>18 years old) patients with OPC diagnoses who were treated with neck RT between June 2016 and June 2019, with a resultant minimum 5-year surveillance interval, at a single tertiary cancer center previously consented and enrolled under a prospective cohort registry [MD Anderson Institutional Review Board protocol PA14-0947] were retrospectively identified and data extracted for inclusion in this study [MD Anderson Institutional Review Board protocol PA2025-0187]. Considering the long latent period between RT and development of clinically evident vascular disease, we selected a study period that would allow for at least five years of follow up and also reflect the modern treatment strategies for OPC (i.e. intensity modulated photon or proton radiotherapy). The cohort was selected from an extant, prospectively maintained registry of consecutive patients with OPC. Patients who had undergone prior neck RT, RT treatment dates were not available or who died during their treatment period of cancer were excluded from this study.

### Clinical Variable Definition

Demographic data as well as baseline clinical and oncological data including RT treatment dates, RT modality, laterality, cancer site and staging, possible surgeries, chemotherapy, and immunotherapy were prospectively collected. To account for dosimetric and radiobiological differences in delivered treatment, carotid-specific iso-effective Equivalent Dose in 2 Gy Fractions (EQD2) values were calculated using the linear-quadratic model applied to collected radiation dose data.^13^ α/β ratios of 2 and 3 have been used in various studies for late-responding normal tissues such as the carotid artery, and we opted for 3 as a more conservative value.^14,15^

Baseline cardiovascular comorbidities and ABF-associated clinical outcomes were abstracted by manual review of individual electronic medical records. Cardiovascular comorbidities included history of hypertension, dyslipidemia (DLD), diabetes mellitus (DM), current or former use of tobacco and obesity (BMI higher than 30 kg/m^2^). Obstructive coronary artery disease (CAD), sustained arrhythmias that required treatment, syncope, TIA or stroke, carotid artery disease (with more than 50% stenosis), peripheral artery disease (PAD), cardiomyopathy or heart failure (HF) and valvular disease (at least moderate) were also captured along with use of cardiovascular medications (statins and antiplatelet agents) before RT.

ABF-associated clinical manifestations that were recorded included new or worsening hypertension, new hypotension, sustained arrhythmias, and syncope. Hypertension was defined as a new increase in the patient’s blood pressure requiring antihypertensive treatment or need for up-titration of baseline antihypertensive therapy. Sustained arrhythmias were captured only if they required intervention. New hypotension was included only when it required pharmacologic intervention (e.g., midodrine, fludrocortisone), excluding cases managed solely with intravenous fluids. Weight changes were recorded for patients who developed hypertension or hypotension to assess whether they contributed to the development of new blood pressure clinical events. Additional endpoints included new or worsening of carotid artery atherosclerosis (including wall thickening, plaque or calcifications) and/or stenosis (more than 50%) on available imaging, TIA, stroke, as well as all-cause mortality. Worsened atherosclerosis was determined based on the interpretation of imaging studies by the radiologist involved in the routine clinical care of the patients. For all clinical outcomes, the date of the initial event was also recorded.

### Statistical Analysis

Demographic and clinical characteristics were summarized using descriptive statistics, with means ± standard deviations (SD) or medians with interquartile ranges (IQR) for continuous variables, and counts (%) for categorical variables. The composite of ABF-associated outcomes including hypertension, hypotension, arrhythmia, and syncope was also summarized by counts (%). Paired t-tests compared baseline weights with weights during hypertension and hypotension events. Two group means were compared using a Welch’s two sample t-test. Univariate and multivariable cause-specific Cox regression models were fitted to identify the covariates that were independently associated with the risk of developing any ABF-associated outcome. Those without an event were censored at the time of death or last follow-up. Proportional hazards assumptions were checked by visual inspection of Kaplan-Meier plots stratified by subgroup. To assess the robustness of the findings, Fine-Gray models were conducted as a sensitivity analyses. These models evaluated the impact of covariates on the cumulative incidence of ABF-associated outcomes following radiation therapy, treating death without an ABF-associated outcome as a competing risk. Aalen-Johansen cumulative incidence plots were generated based on significant covariates identified by univariate models. An *α* < 0.05 was considered significant. Analyses were conducted using SAS 9.4 (SAS Institute Inc., Cary, NC).

## Results

### Cohort Characteristics

A total of 393 patients were identified from within the primary cohort for data extraction (Figure 1). Mean age at diagnosis was 61 ± 10 years, 88% were male, and 91% White (Table 1). Median body mass index (BMI) was 28.9 kg/m² (IQR, 26.3–31.9). Cardiovascular risk factors were present in 87% of the cohort. Former or current use of tobacco products was reported in 55% of patients, hypertension in 51%, DLD in 46%, DM in 16%, and obesity in 29%. A history of cardiovascular disease was present in 23% of patients, including sustained arrhythmias (8%), moderate-to-severe valvular disease (2%), and carotid artery disease with >50% stenosis (4%).

**Figure 1:**
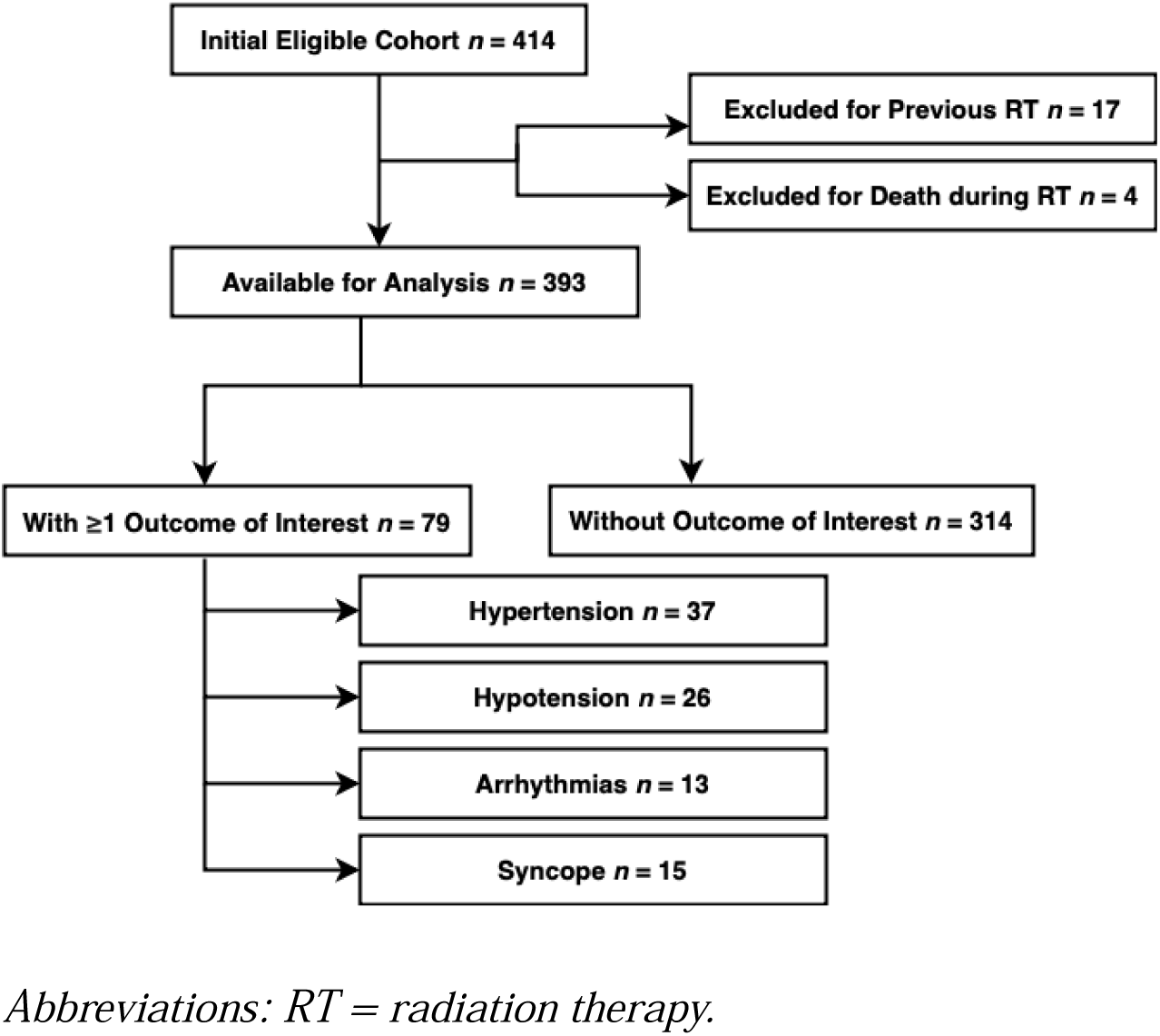
Study Flowchart of Patient Selection.

**Table 1:** Baseline Demographic and Clinical Characteristics of Oropharyngeal Cancer Survivors treated with Radiation Therapy.

|  | Descriptive Statistics |
| --- | --- |
| Age at RT start date (years) – mean ± SD | 60.5 ± 10.0 |
| Female sex – no. (%) | 48 (12.2) |
| Race – no. (%) |  |
| White | 357 (91.1) |
| Black | 9 (2.3) |
| Other | 26 (6.7) |
| Hispanic ethnicity – no. (%) | 35 (8.9) |
| BMI – median (IQR) | 28.9 (26.3-31.9) |
| Obesity – no. (%) | 113 (28.8) |
| Smoking – no. (%) | 217 (55.2) |
| Hypertension – no. (%) | 200 (50.9) |
| Dyslipidemia – no. (%) | 179 (45.5) |
| Diabetes Mellitus – no. (%) | 61 (15.5) |
| Obstructive coronary artery disease – no. (%) | 32 (8.1) |
| Arrhythmias – no. (%) | 32 (8.1) |
| Cerebrovascular accident/TIA – no. (%) | 18 (4.6) |
| Carotid artery disease (>50% stenosis) – no. (%) | 14 (3.6) |
| Peripheral artery disease – no. (%) | 12 (3.1) |
| Cardiomyopathy or heart failure – no. (%) | 9 (2.3) |
| Valvular disease (at least moderate) – no. (%) | 9 (2.3) |
| Statins – no. (%) | 135 (34.4) |
| Antiplatelet – no. (%) | 107 (27.2) |
| Positive p16 HPV – no. (%) | 323 (86.6) |
| Equivalent Dose in 2 Gy Fractions – median (IQR) | 70.7 (68.6-71.6) |
| RT modality – no. (%) |  |
| Proton RT modality | 84 (21.4) |
| Photon RT modality | 303 (77.1) |
| Mixed RT modality | 6 (1.5) |
| T stage – no. (%) |  |
| T0-T1 cancer stage | 142 (36.4) |
| T2-T3 cancer stage | 190 (48.7) |
| T4 cancer stage | 58 (14.9) |
| N stage – no. (%) |  |
| N0-N1 cancer stage | 290 (74.2) |
| N2-N3 cancer stage | 101 (25.8) |
| M stage – no. (%) |  |
| M0 cancer stage | 382 (98.5) |
| M1 cancer stage | 6 (1.5) |
| Stage IV cancer – no. (%) | 125 (32.6) |
| Tumor laterality – no. (%) |  |
| Right | 200 (52.4) |
| Left | 171 (44.8) |
| Bilateral | 11 (2.9) |
| Site of tumor – no. (%) |  |
| Base of tongue site of tumor | 171 (43.5) |
| Tonsil site of tumor | 175 (44.5) |
| NOS site of tumor | 30 (7.6) |
| Other site of tumor | 17 (4.3) |
| Surgery – no. (%) | 42 (10.7) |
| Chemotherapy – no. (%) | 336 (85.5) |
| Type of chemotherapy – no. (%) |  |
| No chemotherapy | 57 (14.5) |
| Taxane-only chemotherapy | 2 (0.5) |
| Platinum-only chemotherapy | 158 (40.2) |
| Platinum-taxane chemotherapy | 101 (25.7) |
| Other chemotherapy agents | 75 (19.1) |
| Type of chemotherapy – no. (%) |  |
| Platinum-taxane chemotherapy | 101 (25.7) |
| Other/No chemotherapy | 292 (74.3) |
| Immunotherapy – no. (%) | 108 (27.5) |

*Abbreviations: BMI = body mass index; HPV = human papillomavirus; IQR = interquartile range; NOS = not otherwise specified; RT = radiation therapy; SD = standard deviation; TIA = transient ischemic attack.*
*Note: Values are expressed as mean ± SD for normally distributed variables, median [IQR] for skewed continuous variables, and number (percentage) for categorical variables.*

Regarding cancer-related characteristics, 33% of patients presented with Stage IV disease. Median total RT dose delivered was 70 Gy (IQR, 66–70), with median EQD2 of 70.7 Gy (IQR, 68.6–71.6). Most patients (77%) received photon-based treatment. Chemotherapy was administered in 85%, with 66% receiving platinum-based regimens and 26% also receiving a taxane. Immunotherapy was given to 27% of patients, and 11% underwent surgery as part of their treatment.

### Incidence of ABF-associated Manifestations and Outcomes

During a median actuarial follow-up of 6.3 years (95% CI: 6.2–6.4), 19.1% of patients developed at least one ABF-associated clinical event. New or worsening hypertension was the most frequent (9.4%), followed by hypotension (5.3%), syncope (3.8%), and arrhythmias (3.3%). New or worsening carotid artery atherosclerosis was observed in 38.9% of patients and significant stenosis (>50%) in 7.1%. Stroke or TIA occurred in 2.3% of the cohort (Table 2). All-cause mortality was 21.4%. Although patients who developed hypertension or hypotension experienced post-treatment weight loss, the magnitude of weight change did not differ significantly between the two groups (p = 0.54). Significant mean weight loss was observed in both groups (−7.6 ± 8.8 kg in hypertension; −10.5 ± 19.0 kg in hypotension), but these changes were not sufficient to fully explain the development of blood pressure lability.

**Table 2:** Prevalence of Outcomes of Interest Among Oropharyngeal Cancer Survivors Treated with Radiation Therapy.

|  | N (%) |
| --- | --- |
| Development of $\geq 1$ Afferent Baroreflex Failure-associated Manifestation | 75 (19.1) |
| Hypertension | 37 (9.4) |
| Hypotension | 21 (5.3) |
| Arrhythmias | 13 (3.3) |
| Syncope | 15 (3.8) |
| Stroke or TIA | 9 (2.3) |
| Carotid artery atherosclerosis | 153 (38.9) |
| Carotid artery atherosclerosis with stenosis $>50\%$ | 28 (7.1) |
| Death | 84 (21.4) |

*Abbreviations: TIA = transient ischemic attack.*

A progressive increase in the composite of ABF-associated manifestations was noted over time following RT. The cumulative incidence of any event rose from 7.4% at 1 year to 23.3% at 7 years, with a steeper increase beginning after 3 years post RT (Figure 2). Manifestation-specific cumulative incidence was also analyzed (Supplementary Figure 1).

**Figure 2:**
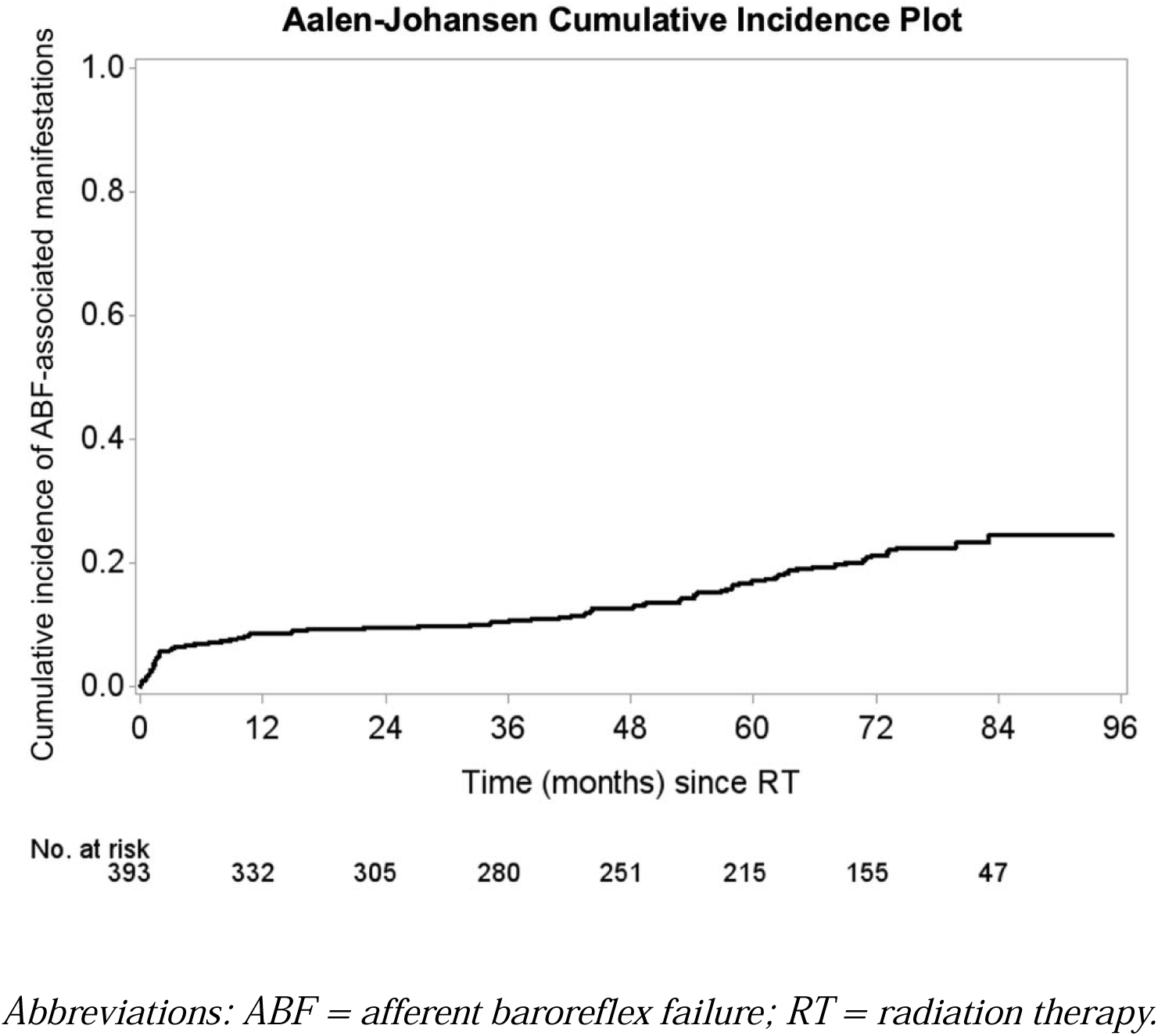
Cumulative Incidence of Afferent Baroreflex Failure-associated Manifestations. This figure illustrates the cumulative incidence of manifestations associated with afferent baroreflex failure following initiation of radiotherapy in a cohort of oropharyngeal cancer survivors. The Aalen-Johansen estimator was used to account for competing risks such as death. Shaded areas represent 95% confidence intervals, and numbers at risk are displayed below the x-axis.

### Clinical Characteristics Associated with ABF Manifestations

In multivariable cause-specific Cox regression analysis, older age (HR 1.03 per year; 95% CI 1.01–1.06), advanced T-category (T4) cancer (HR 1.90; 95% CI 1.10–3.27), moderate or severe valvular disease (HR 2.85; 95% CI 1.03–7.92), and platinum-taxane chemotherapy (HR 1.86; 95% CI 1.13–3.05); were independently associated with an increased risk of the composite of ABF-associated events (Table 4; univariable associations in Table 3). A time-to-event Fine Gray sensitivity analysis was performed accounting for death as a competing risk, and it showed similar findings (Supplementary Table 1 and Supplementary Table 2), supporting the robustness of the findings.

**Table 3:** Evaluation of Clinical Characteristics Associated with Afferent Baroreflex Failure based on a Univariable Cause-Specific Cox Regression Analysis.

| **Covariate** | **HR (95% CI)** | **p-value** |
| --- | --- | --- |
| Age at RT start date (per 1 year) | 1.03 (1.01-1.06) | 0.009 |
| EQD2 (per 1 unit) | 1.00 (0.95-1.06) | 0.995 |
| Male sex vs. Female | 1.36 (0.62-2.95) | 0.443 |
| Race |  |  |
| Black vs. White | 0.66 (0.09-4.73) | 0.676 |
| Other vs. White | 1.66 (0.76-3.63) | 0.200 |
| Not Hispanic ethnicity vs. Hispanic | 0.84 (0.41-1.76) | 0.652 |
| BMI (per 1 unit) | 1.00 (0.95-1.05) | 0.958 |
| Obesity vs. No | 0.93 (0.56-1.55) | 0.787 |
| Smoking vs. No | 0.87 (0.55-1.37) | 0.545 |
| Hypertension vs. No | 1.51 (0.95-2.39) | 0.080 |
| Dyslipidemia vs. No | 1.27 (0.81-2.00) | 0.294 |
| Diabetes Mellitus vs. No | 1.27 (0.68-2.35) | 0.451 |
| Obstructive coronary artery disease vs. No | 1.26 (0.55-2.91) | 0.583 |
| Arrhythmias vs. No | 2.02 (0.97-4.21) | 0.062 |
| Cerebrovascular accident/TIA vs. No | 0.64 (0.16-2.61) | 0.536 |
| Carotid artery disease (>50% stenosis) vs. No | 1.98 (0.72-5.43) | 0.185 |
| Peripheral artery disease vs. No | 0.27 (0.02-4.38) | 0.354 |
| Cardiomyopathy or heart failure vs. No | 0.75 (0.10-5.38) | 0.772 |
| Valvular disease (at least moderate) vs. No | 2.32 (0.85-6.36) | 0.101 |
| Statins vs. No | 1.02 (0.63-1.64) | 0.936 |
| Antiplatelet vs. No | 1.32 (0.81-2.15) | 0.258 |
| Positive p16 HPV vs. No | 0.73 (0.36-1.47) | 0.377 |
| Total RT dose ($\geq 70$ Gy) vs. $\leq 60$ Gy | 0.91 (0.36-2.32) | 0.849 |
| RT modality |  |  |
| Photon RT modality vs. Proton | 1.11 (0.63-1.96) | 0.725 |
| Mixed RT modality vs. Proton | 5.14 (1.48-17.84) | 0.010 |
| T stage |  |  |
| T2-T3 cancer stage vs. T0-T1 | 0.84 (0.49-1.41) | 0.501 |
| T4 cancer stage vs. T0-T1 | 2.08 (1.14-3.78) | 0.016 |
| N2-N3 cancer stage vs. N0-N1 | 1.42 (0.86-2.34) | 0.166 |
| M1 cancer stage vs. M0 | 0.92 (0.13-6.65) | 0.937 |
| Stage IV cancer vs. Non-Stage IV | 1.85 (1.16-2.95) | 0.010 |
| Right tumor laterality vs. left | 0.74 (0.46-1.19) | 0.217 |
| Site of Tumor |  |  |
| Tonsil site of tumor vs. Base of tongue | 0.82 (0.20-3.43) | 0.788 |
| NOS site of tumor vs. Base of tongue | 1.12 (0.47-2.68) | 0.801 |
| Other site of tumor vs. Base of tongue | 1.13 (0.70-1.83) | 0.615 |
| Surgery vs. No | 1.00 (0.50-2.00) | 0.997 |
| Chemotherapy vs. No | 1.18 (0.61-2.30) | 0.622 |
| Platinum-taxane chemotherapy vs. Other/No chemotherapy | 1.95 (1.21-3.15) | 0.006 |
| Immunotherapy vs. No | 1.32 (0.81-2.14) | 0.269 |

*Abbreviations: BMI = body mass index; CI = confidence interval; HR = hazard ratio; HPV = human papillomavirus; IQR = interquartile range; NOS = not otherwise specified; RT = radiation therapy; SD = standard deviation; TIA = transient ischemic attack.*

**Table 4:** Evaluation of Clinical Characteristics Associated with Afferent Baroreflex Failure based on a Multivariable Cause-Specific Cox Regression Analysis.

| **Covariate** | **HR (95% CI)** | **p-value** |
| --- | --- | --- |
| Age at RT start date (years) (per 1 year) | 1.03 (1.01-1.06) | 0.014 |
| Valvular disease (at least moderate) vs. No | 2.85 (1.03-7.92) | 0.044 |
| Stage T4 cancer vs. T0-T3 | 1.90 (1.10-3.27) | 0.021 |
| Platinum-taxane chemotherapy vs. Other/ No chemotherapy | 1.86 (1.13-3.05) | 0.015 |

*Abbreviations: CI = confidence interval; HR = hazard ratio; RT = radiation therapy.*

## Discussion

In this large retrospective cohort study of long-term OPC survivors, nearly one in five developed at least one afferent baroreflex failure (ABF)-associated manifestation—including hypertension, hypotension, arrhythmias, or syncope—after RT. Temporal incidence analysis revealed a notable increase of these manifestations beyond three years from RT. Multivariable cause-specific Cox regression analysis identified older age at diagnosis, advanced T-category (T4) cancer, moderate or greater valvular heart disease, and platinum-taxane chemotherapy as independent risk factors for ABF-related manifestations.

ABF is an underrecognized condition, largely due to its delayed onset—often emerging years after radiation therapy—and its broad, nonspecific clinical manifestations.^16^ Diagnostic difficulty is compounded by overlap with other cardiovascular disorders, leading to frequent misattribution or missed diagnoses.^17^ As a result of these challenges and limited clinical awareness, the true prevalence of ABF remains unknown.^3^ Previous small studies reported variable frequencies of ABF features post-RT or neck surgery, but long-term, large-scale data quantifying cumulative risk in oncologic cohorts have been scarce.^5^ Our study is a start to addressing this gap, estimating that roughly one in five RT-treated OPC long-term survivors experience clinical manifestations that could be related to ABF over a median follow-up exceeding six years. Hypertension was most frequent, followed by hypotension, syncope, and arrhythmias, reflecting the syndrome’s multifaceted nature.^18^

Furthermore, the temporal patterns observed in the development of ABF-related manifestations have important clinical implications for both monitoring and management. While prior studies have identified autonomic dysfunction following radiation therapy, few have characterized its timing or progression.^18^ Our findings demonstrate that ABF-related manifestations most commonly emerge several years after RT, although a subset of patients develops symptoms within the first one to two years. These insights underscore the need for both early and long-term cardiovascular autonomic surveillance in at-risk OPC survivors. Large-scale longitudinal cohort studies are necessary to determine the actual incidence, especially given the excellent long-term outcomes from cancer therapy in OPC populations.^19,20^

Currently, no standardized guidelines exist for cardiovascular autonomic testing or surveillance after RT, which likely contributes to under recognition and diagnostic delays.^10^ Importantly, our retrospective analysis demonstrates a substantial risk (∼20%) of consequential and/or symptomatic ABF, in the absence of coordinated and rigorous cardiovascular surveillance. Emerging data from our group and other suggest surrogate endpoints (such as carotid imaging with US and CT) can detect early sub-acute changes indicative of early direct vascular injury.^25,26^ Similarly, emerging data from our group suggests late cranial neuropathies may manifest during similar time-frames;^27^ together, these data suggest focused survivorship programs capable of monitoring and identifying patients at risk of neurovascular sequelae represents an unmet need, as well as the need for early identification of at-risk populations for ABF, for whom either prophylactic or early mitigation strategies would be ideal.

Patient risk stratification is key towards the development of targeted surveillance and implementation of preventive strategies as well as research into its treatment. Identification of clinical factors that independently predict the development of ABF will assist towards that direction. In our study, we examined the associations of clinical variables with manifestations linked to ABF and we found that older age, advanced T-category (T4) cancer, platinum-taxane chemotherapy and prior moderate-severe valvular disease were independent predictors. Older age likely reflects reduced autonomic reserve, with known age-related declines in baroreceptor sensitivity and heart rate variability, although worsening or incident hypertension, independent of ABF is also associated with increasing age.^18^ Advanced T-category (T4) cancer may indicate more extensive tumor burden and aggressive treatment, increasing nerve injury risk or systemic factors impairing autonomic regulation.^21^ Platinum-taxane chemotherapy’s neurotoxicity is well described and likely synergizes with RT-induced nerve damage to promote ABF.^22^, Moderate or greater valvular heart disease likely contributes to a reduced hemodynamic reserve which make changes in blood pressure and heart rate more evident.^24^ Overall though, the reported associations should not be interpreted as definitive predictors of ABF but rather as clinical variables that may identify patients with reduced autonomic reserve or increased susceptibility to dysregulation. These variables should prompt clinicians to consider autonomic evaluation in appropriate patients.

Radiation dose was not significantly associated with ABF risk in our cohort, which may be due to the narrow dose range centered around high therapeutic levels in patients with OPC, limiting dose-response detection.^21^ Additionally, aspirin and statins did not show protective effects against ABF manifestations but that should interpreted in the context of limitations associated with our retrospective design and cohort characteristics, given that these medications may be used at baseline in higher risk patients. Future prospective studies incorporating detailed autonomic function assessments are needed to clarify these relationships and explore potential preventive strategies targeting ABF in cancer survivors.

### Limitations

This study has inherent limitations related to its retrospective design, including potential selection and information biases from electronic medical record review. For the purpose of this manuscript, we evaluated prevalence of manifestations associated with ABF which may be nonspecific. Our intention was to identify patients who will benefit from dedicated autonomic testing. The predominantly male and White patient population from a single tertiary center limits generalizability. Although follow-up exceeded six years, longer surveillance may be needed to fully capture late-onset events. Unmeasured confounders related to comorbidities or treatment variations may have influenced outcomes despite multivariable adjustment. Moreover, ABF pathophysiology is heterogeneous, suggesting the identified risk factors represent overlapping but distinct mechanisms, restricting causal inference. Future prospective research is needed to clarify biological pathways, validate risk stratification, and guide preventive strategies in OPC survivors.

Despite its limitations, our study, representing, to our knowledge, the largest extant OPC-specific long-term (>5-year) head and neck cohort survey of ABF prevalence is valuable in systematically assessing the prevalence, clinical manifestations, and predictors of this orphan cadiooncology disease state in modern RT-treated OPC survivors.

## Conclusions

In the modern RT era, one in five OPC survivors develop manifestations associated with ABF. While not pathognomonic, these signs warrant evaluation for autonomic dysfunction. Enhanced clinician awareness and patient risk stratification based on comorbidities may facilitate earlier diagnosis and guide future studies on preventive interventions.

## Data Availability

All data produced in the present work are contained in the manuscript. The original dataset used for analysis can be found in an NIH-supported generalist scientific data repository (figshare) at https://doi.org/10.6084/m9.figshare.30081076.v2.

## Data sharing statement

In accordance with NOT-OD-21-013, *Final NIH Policy for Data Management and Sharing*, anonymized/de-identified data that support the findings of this study are openly available in an NIH-supported generalist scientific data repository (figshare) at https://doi.org/10.6084/m9.figshare.30081076.v2 no later than the time of an associated publication.

## Public access policy compliance

In accordance with NOT-OD-25-049, *Supplemental Guidance to the 2024 NIH Public Access Policy: Government Use License and Rights:* “This manuscript is the result of funding in whole or in part by the National Institutes of Health (NIH). It is subject to the NIH Public Access Policy. Through acceptance of this federal funding, NIH has been given a right to make this manuscript publicly available in PubMed Central upon the Official Date of Publication, as defined by NIH.”

## Pre-print Statement

Consistent with NOT-OD-17-050, *Reporting Preprints and Other Interim Research Products*, as “NIH encourages investigators to use interim research products, such as preprints, to speed the dissemination and enhance the rigor of their work”, a pre-peer reviewed deposition of the initial submission version of the manuscript will be available for public access.

## Reporting Guideline Compliance Statement

In accordance with EQUATOR Network (Enhancing the QUAlity and Transparency Of health Research) guidance, we have utilized the RECORD (REporting of studies Conducted using Observational Routinely-collected health Data) checklist; the completed RECORD checklist is attached as Supplementary Table 3 in the Supplementary Appendix.

## CRediT statement

In accordance with the Contributor Roles Taxonomy (CRediT, https://credit.niso.org/), the contributing authors have designated responsibilities and individual author attribution. The corresponding author(s) (EK) assume(s) responsibility for role assignment, and all contributors have been given the opportunity to review and confirm assigned roles.

**Patrik J. Van-Bergen**: Conceptualization, Data curation, Investigation, Methodology, Visualization, Writing – original draft; **Dhawal S. Bandrey**: Data curation, Investigation, Writing – review & editing; **Juhee Song**: Formal analysis, Writing – review & editing; **Stephen P. Patin**: Investigation, Writing – review & editing; **Neeharika Prathapa**: Investigation, Writing – review & editing; **Gabrielle N. Mbagwu**: Investigation, Writing – review & editing; **Andres Hughes**: Investigation, Writing – review & editing; **Jennifer Shen**: Data curation, Writing – review & editing; **Mohamed A. Naser**: Funding acquisition, Resources, Writing – review & editing; **Katherine A. Hutcheson**: Resources, Writing – review & editing; **David I. Rosenthal**: Resources, Writing – review & editing; **Amy C. Moreno**: Funding acquisition, Resources, Writing – review & editing; **Elie Mouhayar**: Resources, Writing – review & editing; **Anita Deswal**: Resources, Writing – review & editing; **Clifton D. Fuller**: Funding acquisition, Resources, Writing – review & editing; **Efstratios Koutroumpakis**: Conceptualization, Methodology, Project administration, Resources, Supervision, Writing – review & editing.

## Abbreviations

ABF: Afferent Baroreflex Failure
CAD: Coronary Artery Disease
DLD: Dyslipidemia
DM: Diabetes Mellitus
EQD2: Equivalent Dose in 2 Gy Fractions
HF: Heart Failure
OPC: Oropharyngeal Cancer
PAD: Peripheral Artery Disease
RT: Radiation Therapy
TIA: Transient Ischemic Attack

---

**Central Illustration:**
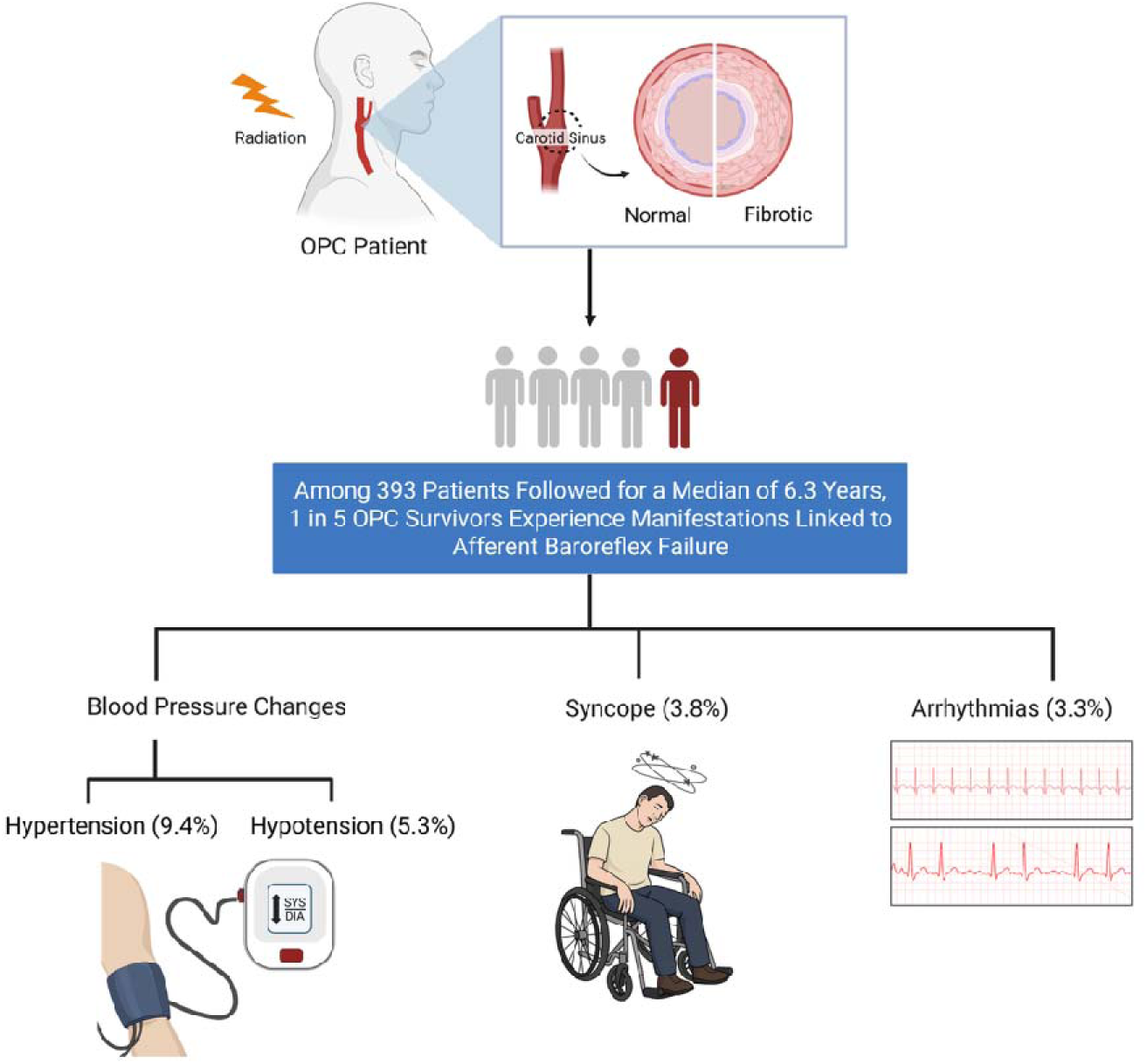
Afferent Baroreflex Failure-associated Manifestations in Oropharyngeal Cancer Survivors Treated with Radiation Therapy. Among 393 patients, during a median follow up of 6.3 years, approximately 1 in 5 developed ABF-associated manifestations, including hypertension (9.4%), hypotension (5.3%), syncope (3.8%), and arrhythmias (3.3%). These findings emphasize the clinical relevance of ABF in long-term OPC survivorship, warranting routine cardiovascular monitoring and potentially informing future screening strategies in this high-risk population.

